# CLINICAL IMPLEMENTATION OF PREEMPTIVE PHARMACOGENOMICS TESTING FOR PERSONALIZED MEDICINE AT AN ACADEMIC MEDICAL CENTER

**DOI:** 10.1101/2024.05.06.24306958

**Authors:** Bani Tamraz, Jaekyu Shin, Raman Khanna, Jessica Van Ziffle, Susan Knowles, Susan Stregowski, Eunice Wan, Rajesh Kamath, Christopher Collins, Choeying Phunsur, Benjamin Tsai, Patsy Kong, Clari Calanoc, Aleta Pollard, Rajeev Sawhney, Jennifer Pleiman, Walter Patrick Devine, Rhiannon Croci, Aparna Sashikanth, Lisa Kroon, Russ Cucina, Aleks Rajkovic

## Abstract

**Objective:** This paper describes the implementation of preemptive clinical pharmacogenomics (PGx) testing linked to an automated electronic clinical decision support (CDS) system delivering clinically actionable PGx information to clinicians at the point of care at UCSF Health, a large Academic Medical Center.

**Methods:** A multidisciplinary team developed the strategic vision for the PGx program. The drug-gene interactions of interest were compiled, and actionable alleles were identified. A genotyping platform was selected and validated at the in-house laboratory. Following HIPAA protocols, genotype results were electronically transferred and stored in EPIC. CDS was developed and integrated with electronic prescribing.

**Results:** We developed a customized clinical PGx program for 56 medications and 15 genes. 233 MWs and 15 BPAs, approved by clinicians, were built into EPIC to deliver actionable clinical PGx information to clinicians.

**Conclusions:** Our multidisciplinary team successfully implemented preemptive PGx testing linked to point-of-care electronic CDS to guide clinicians with precise medication decision-making.

## Background

University of California San Francisco (UCSF) Health recognizes precision medicine (PM) as a priority, with pharmacogenomics (PGx) as an essential PM component. PGx provides a means of tailoring medication treatment to individual patient’s genetics. PGx clinical recommendations are available for many drugs from different resources, including the FDA[1] and the Clinical Pharmacogenetics Implementation Consortium (CPIC)[2]. Despite their availability, PGx-guided therapy is not routinely incorporated into most clinical practices. Many factors currently limit the integration of PGx into clinical care; these can be categorized as bioinformatics and non-bioinformatics.

Bioinformatics factors can be divided into intrinsic and extrinsic categories. Intrinsic challenges involve effectively integrating PGx into clinical workflows within a single electronic health record (EHR) to provide evidence-based, timely, and patient-specific information. Lack of clinical decision support (CDS) in an EHR is a barrier to adopting PGx testing.[3] Solutions are needed to report PGx test results in a standardized, structured, and coded data format for both provider and patient-facing (rather than non-computable PDF and free text formats). Additionally, tools are required to integrate codified PGx test results from multiple sources (e.g., laboratory, CPIC) for efficient clinician access at the point of care. Even if an institution solves the intrinsic challenges related to PGx implementation, these solutions are not easily extended to other EHR systems due to extrinsic factors like interoperability.

Non-bioinformatics factors limiting the broad clinical application of PGx include limited data on clinical utility, long turnaround times for results, limited cost-effectiveness and outcomes data, payer restrictions, lack of consensus guidelines for interpretation, and limited clinician awareness and knowledge.[2 4-7]

Despite challenges, some institutions have integrated EHR-based PGx CDS systems.[8-13] We detail implementing the PGx system at UCSF Health, a large academic health system.

## Methods

### Assemble a multidisciplinary team

A 23-member team with broad expertise in bio-informatics, hospital operations, PGx, EHR usability, software engineers, laboratory operation, clinical education, billing, and clinicians (i.e., physicians, nurses, pharmacists) was assembled and tasked by hospital leadership (medical information, genomics, pharmacy) to investigating PGx implementation specifics, develop a lab process, assess feasibility, and devise clinician education strategies to integrate PGx into UCSF Health’s EPIC EHR system (known as APeX). The team identified the following steps.

### Identify drug-genes interactions of interest

CPIC[2] and FDA[1] table of pharmacogenomic biomarkers were the primary resource for assembling a list of medications and genes. From CPIC, we focused on medications with the highest level of evidence (i.e., level A and B recommendations). We identified medications for which the FDA recommends germline PGx testing before treatment initiation. Then, following consultation with clinical specialists who routinely use the target medications, the list was finalized for program launch.

### Engage clinical leadership for PGx application and implementation across services

Identifying and engaging clinical leadership early in the project was crucial for successfully implementing and adopting PGx. Involving leaders from diverse service lines such as pharmacy, nursing, primary care, and clinical specialties provided a comprehensive understanding of PGx’s potential impact. Clinical leaders championed the integration of PGx into workflows, advocated for resource allocation, and provided guidance on implementation within their services. Engaging leadership fostered buy-in, encouraged interdisciplinary cooperation, and facilitated PGx adoption.

### Identify clinically actionable variants within target genes

This process involved using the PharmGKB[14] table of genes and variants to meticulously identify and prioritize variants associated with known genetic phenotypes.

### Select a genotyping platform and subsequent data analysis

Given UCSF Health’s clinical and genetics expertise and resources, a key decision was made to conduct the PGx testing in our in-house CLIA (Clinical Laboratory Improvement Amendments of 1988) and CAP (College of American Pathologists) accredited clinical genomics laboratory. We began by selecting a genotyping platform. This process involved systematically evaluating various factors to ensure compatibility with clinical objectives. We defined the scope of the clinical application, considering the number of samples, throughput requirements, budget constraints, and the specific genetic variants of interest. We assessed the technical specifications of available platforms, focusing on accuracy, resolution, coverage, and scalability, as well as customization, flexibility, ease of use, data analysis capabilities, and compatibility with downstream applications. Ultimately, the chosen genotyping platform aligned with the project’s objectives, providing reliable and reproducible results while optimizing efficiency and cost-effectiveness.

### Identify & CLIA validate target genetic variants

Identifying and CLIA-validating target genetic variants was a meticulous process critical for ensuring the accuracy and reliability of genetic testing in clinical settings. The process began with the selection of specific genetic variants of interest and identifying reference samples from the Genetic Testing Reference Materials Coordination Program (Get-RM) (https://www.cdc.gov/labquality/get-rm/index.html). Since not every variant of interest had an available reference sample, only those with available references underwent rigorous validation according to CLIA standards. This involved comprehensive laboratory testing to confirm their presence and verify the testing methodology’s accuracy, precision, and reproducibility across multiple samples. Additionally, it included establishing quality control measures, proficiency testing, and adherence to regulatory guidelines to ensure consistent and reliable results.

### Develop best practice advisories and medication interaction warnings in ApeX

Best practice advisories (BPAs) within APeX were chosen to notify ordering clinicians of potential drug-gene interactions in patients who had not been genotyped, providing an opportunity to order the UCSF PGx test. Medication interaction warnings (MWs) communicated potential drug-gene interaction post-PGx testing. Developing BPAs (pre-test alerts) and MWs (post-test alerts) required a multifaceted approach integrating clinical knowledge, technological expertise, and user-centered design principles. Each alert was designed to provide targeted, timely, actionable PGx guidance integrated into existing clinical workflows and decision points, such as medication ordering. We defined the criteria and logic for triggering each alert and refined them to be context-specific, such as provider type (e.g., oncologist vs. pharmacist). Each alert included instructions for ordering PGx testing or prescribing suggestions based on CPIC and FDA recommendations tailored to the patient’s PGx test results. Continuous monitoring and iterative refinement of alert algorithms were necessary to minimize alert fatigue and optimize clinical utility. We have maintained this monitoring since the program’s launch. By following these steps and engaging stakeholders, we developed robust BPAs and MWs.

### Development and validation of automated clinical decision support (CDS) in ApeX

Developing an automated CDS system involved integrating clinical knowledge, data analytics, and information technology through a systematic process. The EPIC’s Genomics module (From Epic Systems Software Company, Verona, WI) was purchased and used to build the CDS. Initially, we defined the objectives and scope of the CDS system and identified specific clinical scenarios or decision points needing CDS support. We then designed and tested algorithms for interpreting genetic results. Iterative testing, validation, and refinement ensured the accuracy, relevance, and usability of the CDS system in real-world clinical settings. Continuous monitoring and feedback mechanisms allow ongoing optimization and adaptation to evolving clinical needs and evidence.

### Develop patient-specific PGx lab reports

While genetic results were delivered into APeX as discrete lab results, a PDF lab report was designed for ease of sharing with patients. The report presented clear, concise information on identified genetic variants, including their clinical significance and potential implications for drug therapy. It is designed for clinicians and patients, using formatting and non-technical language to enhance understanding.

### Develop educational content for patients and clinicians

Creating educational content for patients and clinicians fosters understanding, empowerment, and informed decision-making. Our multidisciplinary team of healthcare professionals, medical writers, and designers collaborated to ensure accuracy, relevance, and accessibility. For patients, content was created using plain language, a sixth-grade reading level, and multimedia formats to enhance comprehension and engagement. Clinician-focused (e.g., physicians, pharmacists, and nurses) content provided evidence-based information on clinical guidelines and best practices, presented concisely and in a user-friendly manner.

## Results

Over 21 months, our multidisciplinary team developed a customized clinical PGx program for 56 medications and 15 genes (**Table 1**). Basic rules for developing robust BPAs and MWs were devised. Each MW alert message contains three components: the clinical recommendation for a specific drug-gene interaction per CPIC or FDA, the genetic phenotype triggering the alert, and the clinical importance of the interaction. BPA statements have two components: the description of a known drug-gene interaction and its associated clinical consequences and standardized language on turnaround time and treatment initiation without genetics. **Figure 1** provides a sample visual for BPA and MW in APeX.

**Table 1.** List of genes and medications selected for clinical PGx testing at UCSF Health.

| **Genes [N=15]** | **Medications [N=56]** |
| --- | --- |
| CYP2C9 | **Warfarin**, Phenytoin, Celecoxib, Flurbiprofen, Meloxicam, Piroxicam, Ibuprofen, **Siponimod** |
| VKORC1/CYP4F2/GGCX | **Warfarin** |
| CYP2C19 | Trimipramine, Sertraline, Imipramine, Escitalopram, Doxepin, **Clopidogrel**, Clomipramine, Citalopram, Amitriptyline, **Voriconazole**, Pantoprazole, Omeprazole, Lansoprazole, Dexlansoprazole |
| CYP2D6 | Trimipramine, Paroxetine, Imipramine, Nortriptyline, Fluvoxamine, Doxepin, Desipramine, Codeine, Clomipramine, Amitriptyline, Venlafaxine, Tramadol, **Tamoxifen**, Atomoxetine, Ondansetron, **Pitolisant**, Aripiprazole, Risperidone, **Pimozide**, **Tetrabenazine** |
| TPMT/NUDT15 | Thioguanine, Mercaptopurine, **Azathioprine** |
| DPYD | Fluorouracil, Capecitabine |
| CYP3A5 | **Tacrolimus** |
| SLCO1B1/ABCG2/CYP2C9 | **Simvastatin**, Atorvastatin, Fluvastatin, Lovastatin, Pitavastatin, Pravastatin, Rosuvastatin |
| G6PD | **Rasburicase**, **Dapsone**, Septra, Sulfasalazine, **Tafenoquine**, **Primaquine** |
| CYP2B6 | Efavirenz |
| UGT1A1 | Atazanavir, Irinotecan |

Medications in red have best practice advisories that notify a provider that a PGx test can be ordered for the medication.

**Figure 1.**
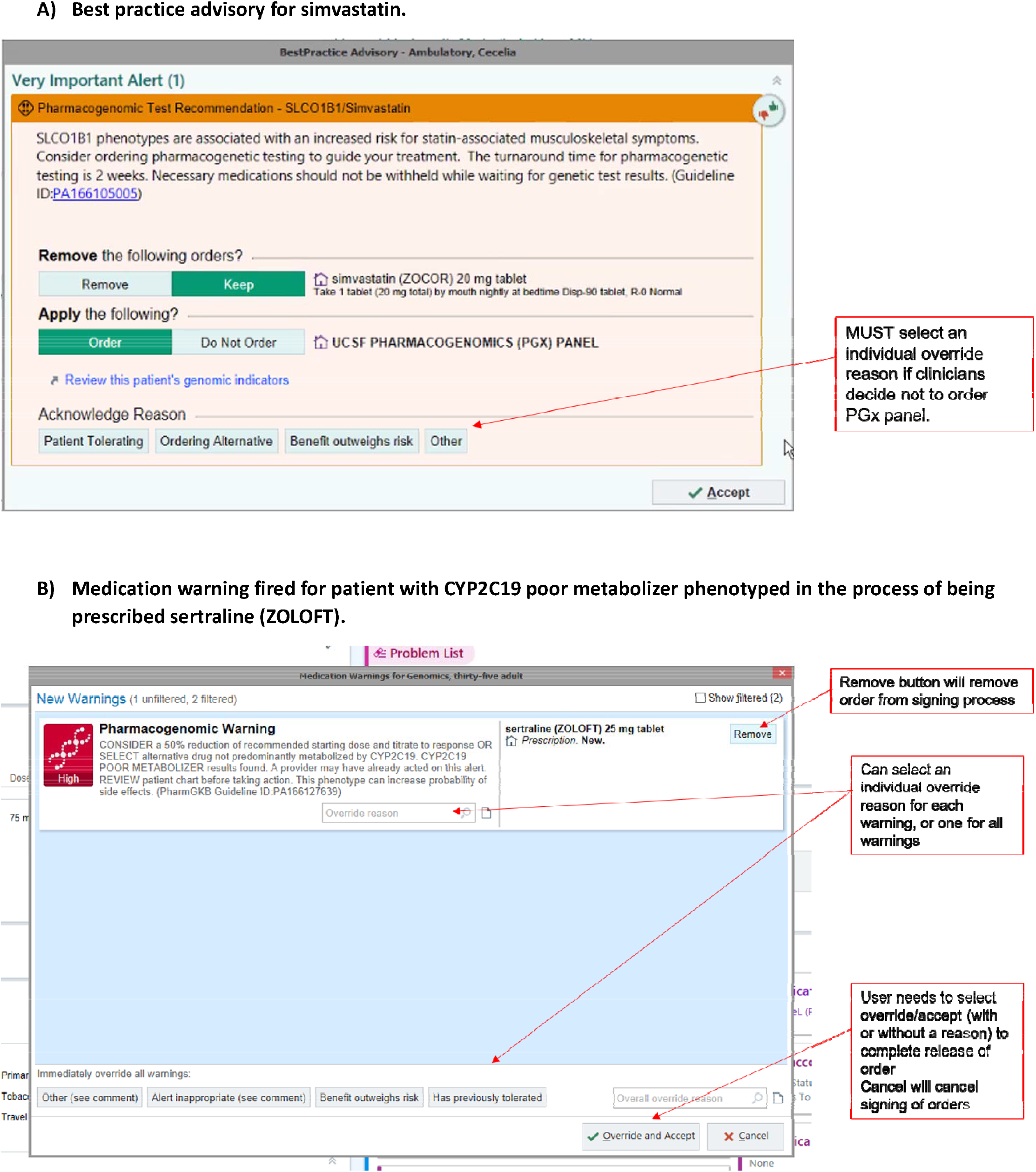
Sample pre-test best practice advisory (A) and post-test medication warning(B)

Based on these rules, 232 MWs and 15 BPAs (**Supplementary File 1**) delivering actionable clinical information to clinicians for 56 medications were drafted, reviewed, and approved by at least 38 clinicians across 16 specialties. These BPAs and MWs were built into APeX’s genomic module and tested using various clinical scenarios. To minimize alert fatigue, the BPAs fire only once per ordering clinician, per patient, and per medication, while the MWs display the information for all actionable drug-gene interactions at appropriate decision points. These 56 medications were prescribed to approximately 120,000 unique patients at UCSF Health in 2020. Given this volume, BPAs were initially implemented for 15 of the 56 medications (**Table 1**). Clinicians can elect not to order PGx testing by selecting one of four acknowledgment reasons displayed in the BPA (**Figure 1**). The UCSF PGx test can be ordered through a BPA presented during medication ordering for 15 medications (if no PGx results exist for the patient) or using the order activity feature within APeX.

The PharmacoScan™ array from ThermoFisher Scientific (Santa Clara, CA) was selected for genotyping target SNPs to construct 240 alleles (**Supplementary Table 1**) with known actionable phenotypes with a 2-week turnaround time. For CLIA validation of the assay, 74 samples (**Supplementary file 2**) were purchased from Coreal. The mean sensitivity and specificity of the assay for identifying the target haplotypes were 99% and 99.5%, respectively.

For ease of use, the translation and integration of structured genetic test results and alerts in APeX are automated. **Figure 2** illustrates the design of a custom pipeline for data flow from ordering the UCSF PGx panel to the resulting, all within APeX. Genomic indicators, essentially phenotypic markers in APeX (e.g., ‘CY2C19 Poor Metabolizer’), are automatically created for the patient based on PGx results from the internal genomics lab. These genomic indicators integrate patients’ PGx results into APeX as structured genetic data that drives the CDS. In addition to discrete genetic results and genomic indicators, a PDF report of the results is generated and placed in the patient’s chart, accessible through Epic System’s MyChart.

**Figure 2.**
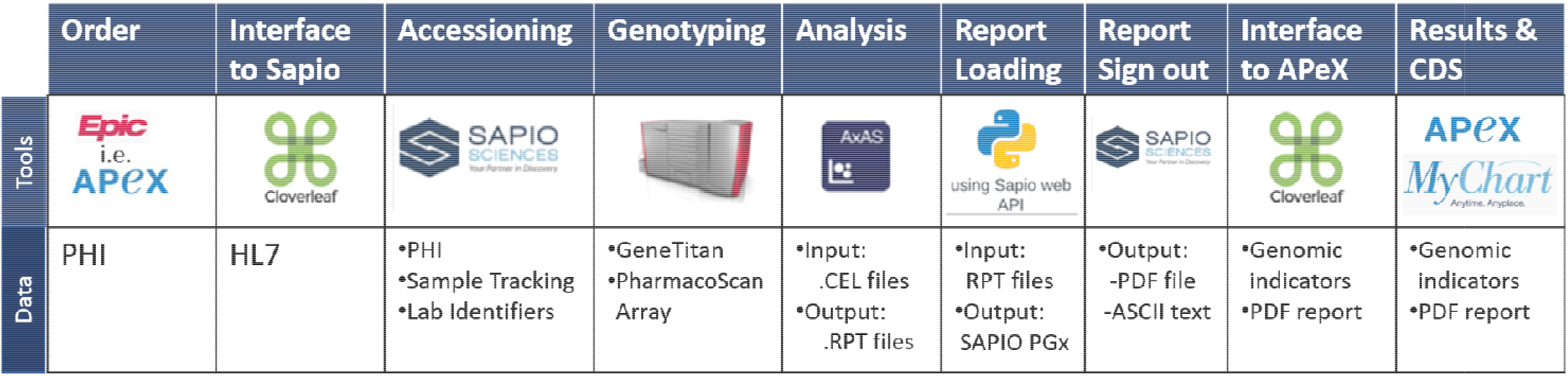
The design of the automated pipeline for the collection of patient information and delivery of structured PGx data to APeX at UCSF Health. Providers order the UCSF PGx test through EPIC^R^ (from Epic Systems Software Company, Verona, WI), known as APeX at UCSF Health. The Cloverleaf^R^ healthcare data platform (From Infor, New York, NY) facilitates the exchange of information between APeX and SAPIO (from Sapio Sciences, Baltimore, MD), the laboratory information management system. UCSF PGx orders are accessioned and tracked in SAPIO. Once a blood or buccal swab sample is received and DNA is extracted, we follow the PharmacoScan™ array protocol (From ThermoFisher Scientific, Santa Clara, CA) for genotyping DNA on the GeneTitan™ instrument from ThermoFisher Scientific. The resulting CEL files generated from GeneTitan™ are important into Axiom™ Analysis Suite software (version 5.1.1.1 from ThermoFisher Scientific) for analysis using customized annotation, metabolizer, and translation libraries. The resulting phenotype.rpt file is scanned for novel alleles and irregularities and uploaded into SAPIO for clinical signout. Once the results are signed out i SAPIO, the associated phenotypes and genotypes are imported back into APeX. Genomic indicators are then automatically created for the patient in APeX, integrating their PGx results into the system through structured genetic data that support clinical decisions. A PDF report of the results is also generated and placed in the patient’s chart, which the patient can access through MyChart from Epic Systems.

All billing is managed through the EHR. The patient’s insurance is billed for PGx testing using the appropriate current procedural terminology (CPT) codes for the medication-gene combination indicated for the PGx test and relevant ICD-10 code(s) associated with the current diagnosis.

Educational content for patients, physicians, pharmacists, and nurses was developed and disseminated. Patients provided verbal consent before ordering the PGx panel.

## Discussion

On May 9^th^, 2023, our multidisciplinary team successfully launched a preemptive PGx testing program integrated with point-of-care automated electronic CDS. This program, which is applied to 56 medications and 15 genes across numerous therapeutic areas, allows us to validate clinical algorithms and assess their efficacy, usefulness, and adoption in routine clinical practice. We envision this project and the developed clinical infrastructure will improve patient health outcomes and provider experience.

The utility of this test will vary across services and clinicians, depending on clinical context, patient factors, and specific medications. By studying the early adopters and provider-patient pairs and considering the range of medications across different therapeutic areas, we expect to gain valuable insights into the implementation processes, including information on reimbursement for PGx testing. Limited information on reimbursement remains a barrier to the widespread adoption of PGx.

As new evidence emerges, our system infrastructure is designed to accommodate additional variants, genes, and medications easily.

## Conclusion

At UCSF Health, we have developed a framework to integrate PGx findings into existing decision-making processes, resulting in more precise, cohesive, and comprehensive recommendations.

## Data Availability

All data produced in the present study are available upon reasonable request to the authors

## Acknowledgements

We acknowledge Joshua Adler, the Chief Medical Officer of UCSF Health, for supporting this project.

## Supplementary material

### Funding

We thank UCSF Health for funding this project.

