## Supplementary table 1 for "CLINICAL IMPLEMENTATION OF PREEMPTIVE PHARMACOGENOMICS TESTING FOR PERSONALIZED MEDICINE AT AN ACADEMIC MEDICAL CENTER"

| **Gene** | **Alleles** |
| --- | --- |
| CYP2C19 | *1,*10,*11,*13,*15,*16,*17,*18,*19,*2,*22,*24,*25,*26,*28,*3,*35, *5,*6,*7,*8,*9,*4.001,*4.002 |
| CYP3A5 | *1, *3, *6, *7 |
| CYP2B6 | *1,*2,*4,*5,*6,*7,*8,*12,*13,*17,*18,*19,*20,*22,*24,*26,*27,*28,*35,*37,*38 |
| CYP2C9 | *1,*2,*3,*4,*5,*6,rs9332094C,*9,*11,*12,*13,*15,*16,*23,*24,*25,*28,*29,*30,*31,*37,*38, *39,*42,*43,*44,*45,*46,*50,*52,*55 |
| CYP2D6 | *1,*2,*3.001,*3.002,*4,*4.009,*4.010,*4.021,*5,*6,*7,*8,*9,*10,*11,*12,*14,*15,*17,*18,*19,*20, *29,*31,*33,*34,*35,*38,*40,*41,*42,*44,*45,*46,*47,*48,*49,*51,*53,*54,*55,*56.001,*56.002,*62, *69,*81,*100,*114, *1XN, *2XN, *41XN, *10XN |
| DPYD | c.=,c.1905+1G>A,c.2846A>T |
| NUDT15 | *1,*3 |
| SLCO1B1 | *1,*5,*9,*14,*15,*23,*31,*20,*37 |
| TPMT | *1,*2,*3C,*4,*11,*14,*15,*23,*29 |
| UGT1A1 | *1,*27,*28,*36,*6 |
| G6PD | B,202A_376G_1264G, A, A-(202A_376G), A-(680T_376G), A-(968C_376G), Acrokorinthos  Amazonia, Amsterdam, Ananindeua, Asahi, Asahikawa, Aures, Aveiro, Bao_Loc, Belem, Canton,  Chikugo, Cincinnati, Cleveland_Corum, Coimbra_Shunde, Costanzo, Crispim, Durham, Gaohe,  Guangzhou, Hammersmith, Harilaou, Hechi, Honiara, Ilesha, Kambos, Kamogawa, Kozukata,  Lages, Lagosanto, Liuzhou, Mahidol, Malaga, Mediterranean, Metaponto, Mexico_City, Miaoli,  Minnesota, Mira_d_Aire, Mt_Sinai, Murcia_Oristano, Musashino, Namouru, Nankang, Nanning,  Naone, Nilgiri, North_Dallas, Orissa, Palestrina, Pedoplis-Ckaro, Plymouth, Quing_Yan, Radlowo,  Rignano, Roubaix, Santa_Maria, Santiago, Sao_Borja, Shenzhen, Shinshu, Sibari, Sinnai,  Songklanagarind, Stonybrook, Sunderland, Swansea, Taipei, Toledo, Tsukui, Ube_Konan, Urayasu  Valladolid, Vancouver, Vanua_Lava, Viangchan, Volendam, Wayne, |
| ABCG2 | rs2231142 |
| GGCX | rs11676382 |
| VKORC1 | rs9923231 |
| CYP4F2 | rs2108622 |
| ---- | rs12777823 |

**Supplementary table 1. Alleles and genetic variants tests for target genes at UCSF Health.**
